# Targeting of Th17-related cytokines in patients with Darier Disease

**DOI:** 10.1101/2023.01.12.22283857

**Authors:** Monika Ettinger, Teresa Burner, Anshu Sharma, Yun-Tsan Chang, Angelika Lackner, Pacôme Prompsy, Isabella Pospischil, Judith Traxler, Gerald Wahl, Sabine Altrichter, Yi-Chien Tsai, Suraj R. Varkhande, Leonie C. Schoeftner, Christoph Iselin, Iris K. Gratz, Susanne Kimeswenger, Emmanuella Guenova, Wolfram Hoetzenecker

## Abstract

Darier disease (DD) is a rare, inherited multi-organ disorder associated with mutations in the ATP2A2 gene. DD patients often have skin involvement characterized by malodorous, inflamed skin and recurrent, severe infections. Therapeutic options are limited and inadequate for the long-term management of this chronic disease. Using gene and protein expression profiling assays, we demonstrate enhanced expression of Th-17-related genes and cytokines and increased numbers of Th17 cells in six DD patients. We prove that targeting the IL-23/-17 axis in DD with monoclonal antibodies is an effective and safe therapy for DD patients, leading to significant clinical improvement. As DD is a chronic, relapsing disease, our findings provide new options for the long-term management of skin inflammation in patients with DD.

## Introduction

Darier Disease (DD) or dyskeratosis follicularis is a rare autosomal dominant inherited disorder (prevalence: 1-3:100.000) defined by heterozygous mutations in the ATP2A2 gene encoding the sarcoendoplasmic reticulum Ca2+ ATPase isoform 2 (SERCA2)^1^. In addition to an increased risk of neuropsychiatric disorders, type 1 diabetes, and heart failure, the skin is the most commonly affected organ in DD patients^2–8^. DD manifests as cutaneous keratotic papules and malodorous plaques, often affecting large areas of the body and significantly affecting patients’ quality of life^9^. The disease has a late onset, often around puberty, and a chronic course with exacerbations triggered by sun exposure, heat, friction, or infection, often requiring hospitalization with intravenous treatment (e.g. antibiotic and/or antiviral drugs)^3^. Therapeutic options are limited and inadequate for the long-term management of this chronic disease with recurrent severe bacterial and viral infections of the skin^9,10^. Conventional treatment still relies on the short-term use of topical corticosteroids, antiseptics and systemic antibiotics. Currently, the most effective treatment is systemic retinoids, but their use is limited by side effects^10,11^. The effects of the disease-causing mutations in the ATP2A2 gene are well studied in the epidermis, where they result in aberrant Ca2+ signaling, loss of intercellular connections (acantholysis) and apoptosis of keratinocytes ^12–15^. By contrast, the consequences of ATP2A2 gene mutations in other cell types, particularly immune cells, are poorly understood^16^. Because chronic inflammation of the skin is common and deleterious in DD, we hypothesized that the involvement of the skin’s immune system is important in the pathogenesis of DD and aimed to characterize the cellular and molecular composition of the cutaneous immune infiltrate in DD skin lesions.

## Results

We included six patients with DD in our cohort. DD was verified by histology of skin biopsies and genetic analyses. The characteristics and previous therapies of the patients (3 males, 3 females; age range 21-58 years) are summarized in Table 1. The patients had skin manifestations since adulthood, ranging from moderate (DD patient number 4 [PAT4]) to severe (PAT1, 2, 5, 7, 9) forms of disease with recurrent bacterial and viral cutaneous infections, some of which required hospitalization (Table 1, Suppl. Fig. 1). All patients had been previously treated with various drugs that provided no or only short-term clinical benefit (Table 1).

**Table 1:** Darier Disease patient characteristics and treatments.

| Patient age*/sex | Heterogenous ATP2A2 mutation | Phenotype/ disease severity** | Complications | Prior therapies | Increased cytokine*** | Current therapy |
| --- | --- | --- | --- | --- | --- | --- |
| PAT1/26-30/f | c.215 C > A, (p.Ser72Tyr) | Severe | recurrent bacterial cutaneous superinfections | topical steroids, antiseptics, systemic therapy with isotretinoin 06/18-02/19 (no beneficial effect) | IL23A | systemic therapy with guselkumab for 13 months |
| PAT2/41-45/m | c.1184T>G, (p.Val395Gly) | Severe | recurrent bacterial cutaneous superinfections | topical steroids, antiseptics, systemic therapy with acitretin 08-09/2017, 04/21-09/21 (no benefit), CO<sub>2</sub> laser on legs (temporary benefit) | IL17A | systemic therapy with secukinumab for twelve months |
| PAT4/46-50/m | c.2305G> A (p.Gly769Arg) | Moderate | recurrent HSV infections and bacterial cutaneous infections | topical steroids, antiseptics, systemic therapy with acitretin 12/18-07/19 (no benefit), CO<sub>2</sub> laser on legs (temporary benefit) | IL17A | topical steroids, antiseptics |
| PAT5/21-25/f | c.392G>A p.(Arg131Gln) | Severe | recurrent bacterial cutaneous superinfections | no topical or systemic therapy | IL17A | loss of follow up |
| PAT7/56-60/m | c.224delT p.(Leu75Trpfs*15) | Severe | recurrent bacterial cutaneous superinfections | topical steroids, antiseptics | IL17A | systemic therapy with acitretin since for four months |
| PAT9/45-50/f | c.1287+1delG splice mutation | Severe | recurrent bacterial cutaneous superinfections depression | topical steroids, antiseptics, systemic therapy with isotretinoin 2008-09/22 (temporary benefit) | IL17A | systemic therapy with secukinumab for six months |

\* age in 5-year ranges
\*\* No standardized assessment scale has been validated to date for DD. The disease was considered severe if the patient had a body surface area (BSA) > 35%, moderate if BSA was 10%-35%.
\*\*\* Cytokine that has been shown to be increased in the patient's lesional skin.

Using single cell RNA sequencing (scRNA-seq), we profiled cells from skin biopsies of DD lesions, which we compared with publicly available scRNA-seq data from psoriasis and normal healthy skin (Fig. 1A, Suppl. Fig. 2)^17^. Based on cluster analysis, 13 cell populations with distinct expression profiles were defined (Suppl. Fig. 2). Cell populations were found in all three groups, with increased numbers of T cells in both DD and psoriasis samples consistent with the inflammatory nature of these diseases (Fig. 1A, Suppl. Fig. 2). Gene expression analysis and subsequent gene set enrichment analysis (GSEA) of DD keratinocytes revealed overexpression of genes involved in mTORC1 signaling and p53 pathway when compared with keratinocytes from normal, healthy skin (Suppl. Fig. 3, Suppl. Table 1). Because an important clinical aspect of DD is chronic skin inflammation, we next performed high-throughput immunoprofiling using NanoString technology (Fig. 1B) of genes expressed in the lesional skin of six DD patients. GSEA of the gene expression data revealed that IL-17-signaling was enhanced in the skin of DD patients compared with the skin of healthy controls (HC; Fig. 1B, C, Suppl. Table 2). Interestingly, the expression profile resembled that of patients with psoriasis, which is known to be a Th17 driven disease (Fig. 1B, Suppl. Table 3)^18^. Based on these results, we examined Th17-, Th1-, and Th2-related cytokines in DD patients by qRT-PCR and found significantly increased IL17A expression compared to HC skin (Fig. 1D, Suppl. Fig. 4). Specifically, five of six DD patients showed increased expression of IL17A and one patient displayed increased expression of IL23A (Suppl Fig. 4, Table 1). Enhanced expression of IL-17A and IL-22, which is produced by several populations of immune cells at a site of inflammation, was confirmed at the protein level by bead-based immunoassays of the protein extract of DD skin biopsies compared with HC skin (Fig. 1E).

**Fig 1:**
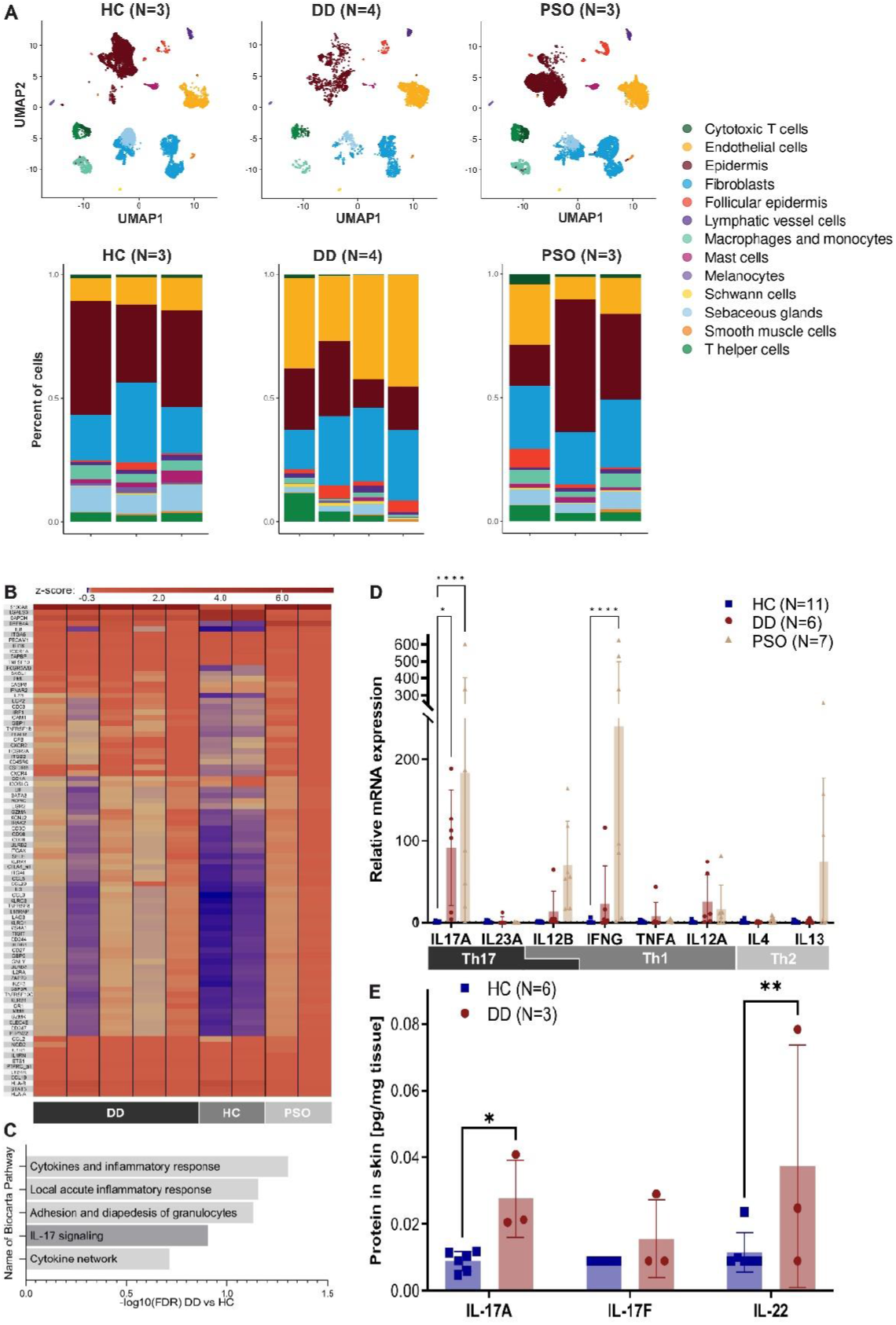
Enhanced expression of Th-17-related genes and cytokines in the skin of patients with Darier disease (DD) **A** UMAP analysis (upper panel) and percentage of different cell types (lower panel) determined by scRNA-seq analysis of skin samples from four DD patients (lesional skin, N=4, 3746 cells). Results were compared to publicly available scRNA-seq data of psoriasis (PSO, lesional skin, N=3, 11417 cells) and three healthy controls (HC, N=3, 12817 cells)^17^. **B** Heat map of differentially expressed (DE) genes expressed as Z-score (DD vs HC, fold change > 1.5) as determined by NanoString nCounter analysis using the Immunology_V2 panel in skin of untreated DD patients (N=5), healthy controls (N=2) and psoriasis patients (N=2). **C** Biocarta pathways enriched in DD patients as compared to HC, as determined by NanoString nCounter and Gene Set Enrichment Analysis. Pathways are ordered according to their normalized enrichment scores. Bars represent -log10 of False Discovery Rates (FDR) (See Suppl Table 2 for complete table). **D** Relative mRNA expression (qRT-PCR) of Th1, Th2 and Th17-associated cytokines normalized to housekeeping gene ACTB and relative to HC. Bars represent means, error bars represent standard deviations. * p<0.05, ** p<0.005, **** p<0.00005 (2-way ANOVA with Dunnett multiple comparison correction (all against HC). **E** Protein expression of Th17-related cytokines in the skin of HC and DD patients as determined by bead-based immunoassays (LegendPlex analysis). * p<0.05, ** p<0.01 (2-way ANOVA).

IL-17(A-F) are pro-inflammatory cytokines produced mainly by a group of T helper (Th) cells known as Th17 cells. Th17 cells play a major role in the pathogenesis of various autoimmune diseases (e.g. rheumatoid arthritis, inflammatory bowel disease, and psoriasis)^19^. To identify the origin of IL-17 in the lesional skin of DD patients, we analyzed IL17 gene expression at the single cell level in our scRNA-seq data set. Among the different cell compartments/types, we detected IL-17A/F expression predominantly in the Th cell subset in skin samples of DD and psoriasis (Fig. 2A) suggesting that Th17 cells are the main source of IL-17 in inflamed skin of patients with DD. The presence of Th17 cells in DD was confirmed by multi-color immunofluorescence (IF) stainings of tissue micro array (TMA) sections (Fig. 2B, Suppl. Fig. 5) and conventional IF stainings of paraffin sections (Suppl. Fig. 6). Quantification of TMA stainings revealed increased numbers of Th17 cells (CD4^+^IL-17^+^) in DD skin samples compared with skin lesions of patients with atopic dermatitis (AD), which is known to be a classical Th2-(IL-4/IL-13) dominated skin disease (Fig. 2C)^20^. In line with these tissue-data, flow-cytometric analysis of PBMC revealed increased numbers of IL-17-producing CD103^+^CLA^+^ circulating skin-resident memory CD4^+^ T cells (cT_RM_) in the blood of patients compared to HC (Fig. 2D). cT_RM_ are cutaneous T_RM_ cells that have exited the skin and can be found in the circulation. In consequence, these cells share cytokine profile with skin T_RM_ and can therefore be utilized to analyze skin T_RM_ from the blood^21^.

**Fig 2:**
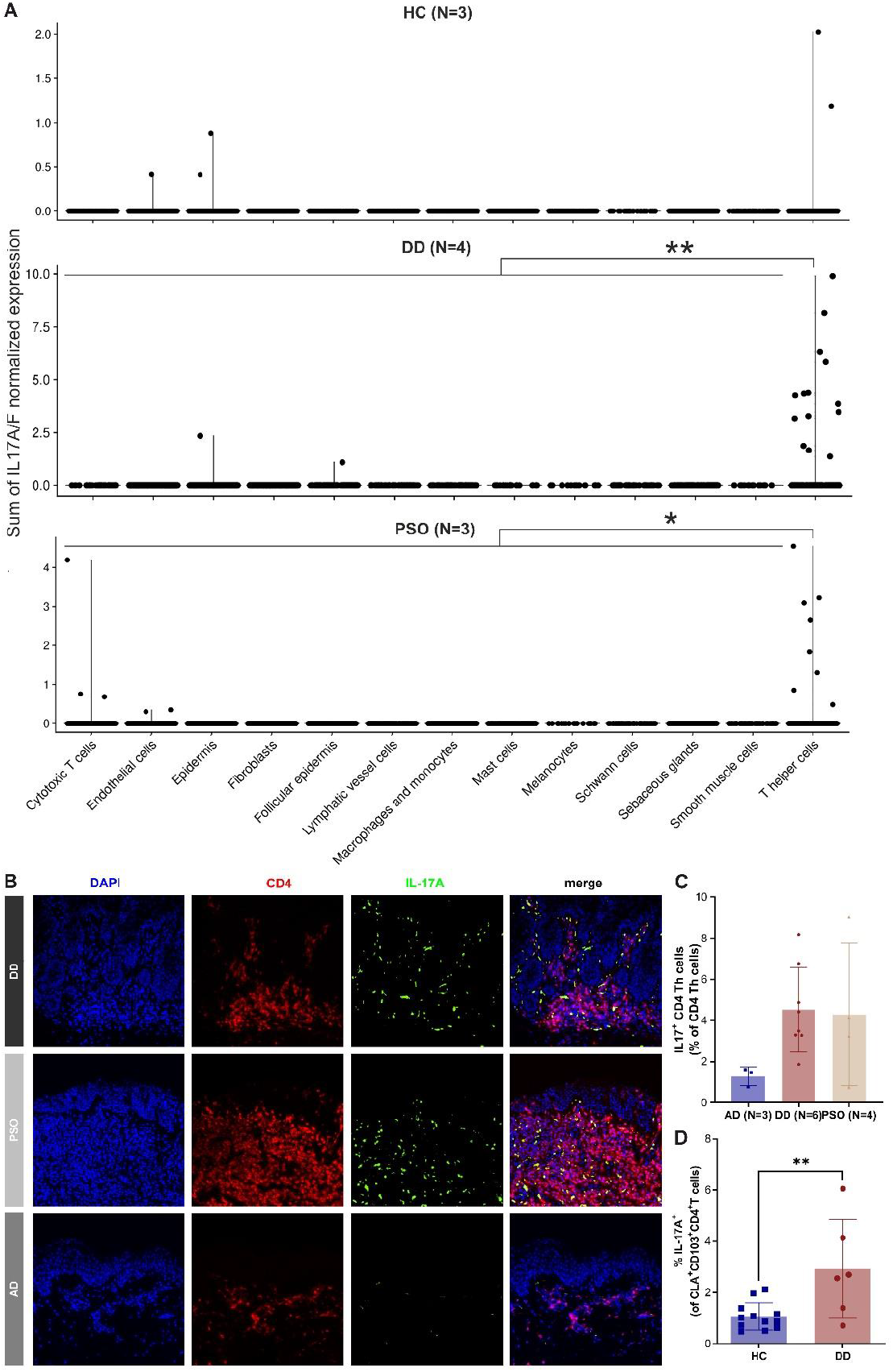
Enhanced numbers of Th17 cells in lesional skin of DD patients. **A** Sum of IL17A/F normalized expression in different cell types HC (N=3, 12817 cells), DD (N=4, 3746 cells) and PSO (N=3, 11417 cells) skin samples as determined by scRNA-seq analysis. Significance bars represent results of student’s T-test of IL17A/F expression between T helper and non-T helper cells, * p<0.01, ** p<0.0005 **B** Representative multi-color immunofluorescence (IF) images of tissue micro array sections stained with OPAL technique for IL17A, CD4 and DAPI in skin samples of DD patients (N=6 patients), atopic dermatitis patients (AD, N=3 patients) and psoriasis patients (PSO, N=4 patients). CD4 positive T cells are marked in red, IL-17A positive cells are marked in green, DAPI-stained nuclei are marked in blue. Isotype controls are shown in Suppl. Fig. 5 **C** Quantification of IL17A/CD4/DAPI triple-positive cells obtained from multi-color immunofluorescence stained TMA sections (skin samples: DD (N=6 patients), PSO (N=4 patients), AD (N=3 patients)). **D** Graphical summary showing IL-17A production by live gated human CD3^+^CD4^+^CD45RA^-^CLA^+^CD103^+^ circulating skin-resident memory T cells (cT_RM_) in the blood of patients compared to HC. Flow cytometric analysis of IL-17A upon ex vivo stimulation with PMA/ionomycin and intracellular cytokine staining.

Because our results indicated an enhanced IL-23/-17 axis in inflamed DD skin, we took a precision medicine approach and administered monoclonal antibodies targeting the overexpressed cytokine to the treatment-refractory patients. Specifically, we treated PAT1 with an IL23A blocking antibody (guselkumab) and PAT2 and PAT9 with an IL17A blocking antibody (secukinumab). In all three patients, we observed a rapid decrease in skin inflammation, followed by a reduction and flattening of hyperkeratotic papules and plaques, especially in the most severely affected areas of the body (i.e., the thorax; Fig. 3A, Suppl. Fig. 7). The assessment of clinical scores showed a 50% reduction of the Investigator’s Global Assessment (IGA) from 4 points (severe disease) at baseline to 2 points (mild disease) during treatment (Fig. 3B). In addition, we observed a reduction in itching and an improvement in the patients’ quality of life (Suppl. Fig. 8). The treatment is well tolerated and has been used in two patients (PAT1, 2) for more than a year. Interestingly, three months after treatment initiation in PAT1 and PAT2, improvement in clinical symptoms was accompanied by normalization of the inflammatory gene expression profile (NanoString and qRT-PCR; Fig. 3C, D) in the patients’ skin. In detail, antibody therapy specifically suppressed Th17 associated cytokines (IL23A in PAT1, IL17A in PAT2) in skin samples, which correlated with a normalization and/or increased expression of Th1 and Th2 cytokines compared to HC (Fig. 3C). The decrease in IL23 expression (Fig. 3C, upper panel) correlated well with a decrease in Th17 cells in PAT1 skin samples collected before and during antibody therapy (Fig. 3E).

**Fig 3:**
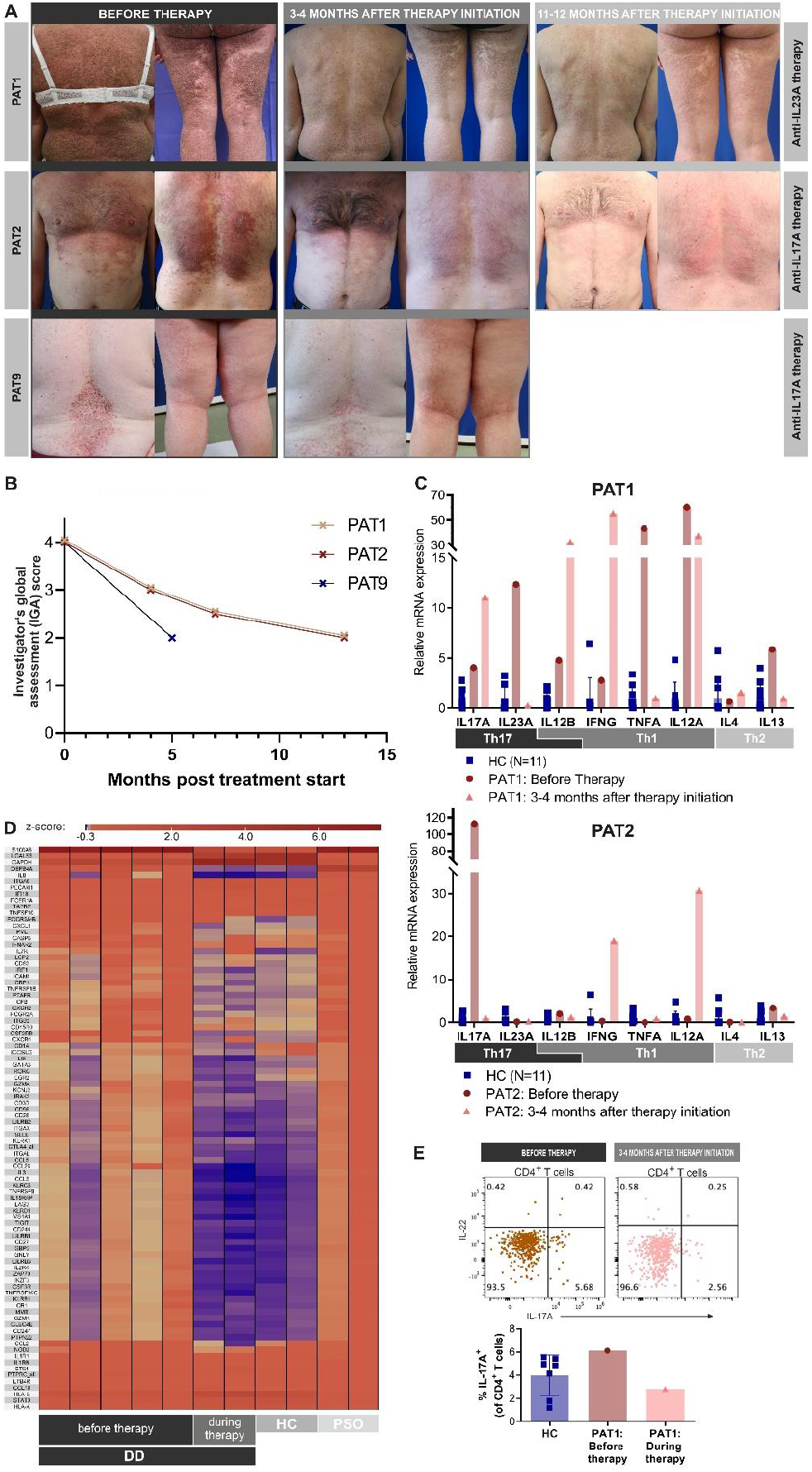
Targeting the IL-23/IL-17 axis with monoclonal antibodies is effective in patients with Darier disease. **A** Cutaneous improvement after personalized targeted antibody therapy. Clinical images of guselkumab (upper, PAT1) and secukinumab (central and lower, PAT2 and PAT9) treated patients before, 3-4 and 11-12 months after initiation of antibody therapy. **B** Clinical improvement of DD patients during therapy as determined by the Investigator’s Global Assessment (IGA) score. **C** Relative mRNA expression (qRT-PCR) of Th1, Th2 and Th17-associated cytokines normalized to housekeeping gene ACTB and relative to HC for PAT1 (upper panel) and PAT2 (lower panel). Bars represent means of different individuals; error bars represent standard deviations. **D** Normalization of gene expression during therapy of differentially regulated genes (DD vs HC) in DD patients determined by NanoString nCounter analysis using the Immunology_V2 panel. Genes represent DE genes expressed as Z-score (DD vs HC, fold change > 1.5). **E** Assessment of Th17 cell counts with multiplex flow cytometry in skin samples from PAT1 before and during therapy with guselkumab.

## Discussion

In our patient cohort, we demonstrated for the first-time increased expression of Th-17-related genes and increased numbers of Th17 cells in the skin of DD patients. Based on our observation, we targeted the IL-23/-17 axis in patients, resulting in an effective and safe therapy.

Treatment of genodermatoses in general is challenging^22^. Because of the small patient population, it is almost impossible to conduct clinical trials and thus get drugs approved. There is a very limited number of effective treatments for DD, and there are no randomized, placebo-controlled trials^9^. Studies addressing the pathophysiology of DD have mainly focused on the epidermis, e.g. keratinocytes. In this context, recent reports have shown that mutations in the ATP2A2 gene lead to impaired calcium homeostasis in keratinocytes, decreased cell-cell adhesion in the epidermis and apoptosis in keratinocytes^3^. This mechanism is thought to lead to the characteristic skin phenotype of DD with hyperkeratotic papules and plaques and provides further rationale for the use of keratolytic retinoids in the long-term management of patients. Indeed, systemic retinoids (e.g. isotretinoin) are effective in DD patients but are often discontinued due to side effects and the persistence of skin inflammation with recurrent infections^11^.

A hallmark of DD is chronic inflammation of the skin, which is common and deleterious in most patients. Interestingly, reports on the role of the immune system in the pathogenesis and progression of DD are rare. To date, skin inflammation in DD patients has mainly been considered an indirect, secondary effect of impaired epidermal barrier function and/or dysbiosis of the cutaneous microbiome in DD patients. In a recent report by Chen et al., mutations in the ATP2A2 gene were for the first time directly associated with impaired immune system function, particularly the B cell compartment^23^. The authors showed that SERCA2, encoded by the ATP2A2 gene, is required for V(D)J recombination and subsequent B cell maturation. Interestingly, some DD patients have reduced numbers of mature B cells in the blood^23^. In our study, we observed an increased number of Th17 cells in the circulation and skin of DD patients, which is comparable to the cell numbers found in psoriasis. On the one hand, the observed Th17 signature in the skin of DD patients could be a direct effect of ATP2A2 gene mutations in immune cells that drives Th17 skewing of naïve T cells. On the other hand, impaired epidermal homeostasis and altered microbiome could be indirectly responsible for increased expression of Th17-related genes and increased numbers of Th17 cells in the skin of DD patients. While psoriasis, as a classic Th17-dominant skin disease, is thought to be triggered at least in part by yet unknown autoantigens and not necessarily by microbial antigens, the altered microbiome of DD skin lesions with recurrent infections may promote chronic inflammation with a Th17 phenotype.

In summary, we demonstrated for the first-time increased expression of Th-17-related genes and enhanced numbers of Th17 cells in DD. We show that targeting the IL-23/-17 axis in DD is an effective and safe therapy in three DD patients. Because DD is a chronic, relapsing disease complicated by recurrent bacterial and viral skin infections, our results provide new options for the long-term management of skin inflammation in patients with DD. Further studies are needed to evaluate the long-term benefits and side effects of this treatment modality and to clarify the formation and role of Th17 cells in DD patients.

## Methods

### Patients and patient samples

#### Molecular genetic analysis

Using Twist comprehensive Exome and Mitochondrial Panel (Twist Bioscience) sequencing on NextSeq2000 (Illumina) as “paired and end reads”.

#### Patient samples

Skin samples (6 mm biopsy specimens) and blood samples for collection of PBMCs were obtained from patients with Darier Disease (DD) (before initiation of therapy and at specific time points during therapy), healthy controls (undergoing cosmetic surgery), and psoriasis patients (PSO) as positive controls. Six mm biopsies were collected on ice in RPMI medium (10.040.CV, Corning), cut into small pieces and slow frozen in FBS (10500064, Gibco) supplemented with 10 % DMSO (D4540, Sigma-Aldrich) within 30 minutes after collection, at the most ^24^. Slow frozen biopsies were used for gene and/or protein expression profiling, as described in the respective sections. PBMCs were isolated using Ficoll-Hypaque (GE Healthcare, GE17-1440-02) gradient separation from the collected blood samples. Isolated PBMCs were cryopreserved using 10% DMSO in FBS and were used for flowcytometry. Written informed consent was obtained from patients and controls. The study was approved by the ethics committee of the Medical Faculty of Johannes Kepler University Linz (EK Nr: 1327/2021, 1036/2020 and 1071/2021) and the ethics committee of the University of Lausanne (CER-VD 2021-00878). The study is registered with ClinicalTrials.gov (NCT05680974).

#### Therapy regimen

Three DD patients (PAT1, PAT2 and PAT9) were treated off-label as part of a precision medicine approach with secukinumab (targeting IL17A, PAT2 and PAT9) or guselkumab (targeting IL23A, PAT1). PAT1 received guselkumab 100 mg subcutaneously at weeks 0 and 4, followed by injections every 8 weeks. PAT2 and PAT9 received secukinumab 300 mg subcutaneously once a week for 5 weeks, followed by monthly administrations.

#### Clinical scores

The Investigator’s Global Assessment (IGA) is a standardized severity assessment in dermatology ranging from 0 (clear) to 4 [severe (deep/dark red erythema, and marked and extensive induration/papulation; excoriation and oozing/crusting present)]. Intermediate scorings were given half points. The DLQI 10 is a 10-item questionnaire assessing health-related quality of life over the last week in patients with dermatological symptoms, with each item scored for impact from not at all (0) to very much (3) resulting in a total score ranging from 0-30 points. Itch was assessed by visual analogue scale ranging from 0-10 points (0 = no pruritus, > 0-< 4 points = mild pruritus, ≥ 4-< 7 points = moderate pruritus, ≥ 7-< 9 points = severe pruritus, and ≥ 9 points = very severe pruritus).

### Gene Expression Profiling

#### RNA extraction

3-mm biopsies of patients’ lesional skin and controls’ normal skin were kept at -80 °C in RNAlater® (Invitrogen). After thawing, biopsies were mechanically disrupted using a TissueLyser II and 5-mm Stainless Steel Beads (Qiagen). RNA was isolated using Monarch Total Miniprep Kit (New England Biolabs) according to the manufacturer’s protocol.

#### Nanostring

For multiplex gene expression profiling of the mRNA, the Nanostring® nCounter System was used together with their immunology panel NS_Immunology_v2_C2328 and the nCounter SPRINT Profiler was used. Positive control was mRNA from skin lesions of two psoriasis patients. Data was analyzed using nSolver 4.0, R and Gene Set Enrichment Analysis (GSEA). ^25,26^ Unfortunately, quality control for PAT2 failed and this patient was excluded from the analysis. Data was nomalized by using all housekeeping genes from the panel with an average count higher than 100 and with less than 35 % coefficient of variance. The list of differentially regulated genes was generated by comparing the expression of all genes between DD and HC samples. Cut-off for fold change was 1.5 and for p-value 0.05 (Student’s t test). Heatmaps were created with the R superheat package. ^26,27^ Data from each gene was scaled to mean 0 and standard deviation 1 (z-score).

#### GSEA analysis

The enrichment score (weighted Kolmogorov-Smirnov-like statistic) tells the degree to which a gene set is overrepresented at the extremes of the ranked list of compared groups (i.e. DD vs HC in Suppl Table 2 and PSO vs HC in Suppl Table 3), normalized enrichment score (NES) accounts for different gene set sizes and correlations between the expression data set and the gene sets. We analyzed the list of gene set ‘Biocarta Pathways’ (c2.cp.biocarta.v2022.1.Hs.symbols.gmt [Curated]. We took a closer look at all gene sets with false discovery rates (FDR) lower than 0.25.

#### Reverse Transcriptase qPCR

cDNA Synthesis Kit (Biozym) was used for cDNA synthesis according to the manufacturer’s instructions. qRT-PCR was conducted on a LightCycler® 450, using DNA Master Fast Star HybProbe (Roche) and Taqman® Gene Expression Assay (FAM) for IL4 (Hs00174122_m1), IL12A (Hs01073447_m1), IL12B (Hs01011518_m1), IL13 (Hs00174379_m1), IL17A (Hs00174383_m1), IL23A (Hs00372324_m1), TNFA (Hs00174128_m1) and INFG (Hs00989291_m1) (Thermo Fisher Scientific). CT values of undetectable genes were set to 45 (lower limit of detection = number of circles recorded in qPCR experiments).

#### scRNA-seq

Slow-frozen skin biopsies were washed with RPMI 1640 medium (Gibco, 61870-010), supplemented with 10% Gold FBS (PAA Laboratories, A15-151). The skin biopsies were then minced and transferred in 1mL of Liberase (Roche, 5401119001, final concentration 0.5 mg/mL diluted with PBS without Ca and Mg) to digest the tissues. The mixture was incubated for 1 hour at 37°C and the tube was manually inverted every 15 minutes. Afterwards, 100 μl of 0.05% trypsin was added to the mixture and incubated at 37°C for additional 15 minutes. The tissue remnants were removed by using 70 μm cell strainer. The cells were washed with MACS buffer (PBS, 0.5% BSA and 2mM EDTA) and then stained with SYTOX Red (ThermoFisher, S34859) and Hoechst 33342 (ThermoFisher, H3570) for cell viability assessment. The stained cells were suspended in MACS buffer and single living cells (SYTOX Red-negative and Hoechst 33342-positive) were sorted by FACS sorter (BD FACSAris-II SORP) at Flow Cytometry Facility of UNIL. Single-cell mRNA capture and sequencing were performed immediately by the Lausanne Genomic Technologies Facility (GTF) of UNIL using the Chromium Next GEM Single Cell 5’ GEM kit (10x Genimics) following the manufacturer’s protocol. Raw sequencing reads were subjected to demultiplexing and aligned to the human reference genome (refdata-gex-GRCh38-2020-A) using the cellranger count tool with default parameters. The analysis included a combination of publicly available dataset (GEO portal number GSE162183) ^17^ comprising psoriasis and healthy control samples, as well as an in-house dataset consisting of four Darier patients. The datasets were processed together by merging the expression matrices based on common genes. To ensure data quality, cells with a read mapping rate of over 25% to mitochondrial genes, which is indicative of dying cells, were filtered out. The remaining cells were then subjected to further analysis using the Seurat package (version 4.3.0) ^28^. Integration of the two datasets was performed using Harmony (version 0.1.1) ^29^, with the retention of the top 50 principal components. Subsequently, cell clusters were identified using the Louvain algorithm implemented in Seurat with a resolution parameter of 0.1. To assign biological annotations, known marker genes were utilized. In addition, the T cell cluster was subjected to additional clustering to distinguish T helper and cytotoxic T cell subsets.

For the differential analysis, Darier and healthy keratinocytes were first pseudo-bulked into 3 randomly assigned replicate each. Then, gene expression was compared and p-values calculated using edge LRT (1) model. P-values were corrected for multiple testing using Benjamini-Hochberg method. Genes were defined as significantly overexpressed if they had a fold-change greater than 2 and a corrected p-value lesser than 0.01. The gene set analysis was run using enrichR (2) on pathways from the following databases: MSigDB_Hallmark_2020, GO_Molecular_Function_2021 and SigDB_Oncogenic_Signatures, retrieving the top pathways with a p-value lesser than 0.1.

### Protein expression profiling

#### Tissue microarray and Opal staining

The skin biopsies from patients were collected and FFPE histology blocks were generated with standard pathology procedures. The tissue microarray (TMA) with different type of skin diseases was created by TMA Grand Master (3DHISTECH). The immunostaining was performed with Opal Manual Detection Kit (NEL861001KT, Akoya Biosciences) according to manufacturer’s instructions. The TMA slide was incubated in two rounds of stainings; in the order of CD4 (ab133616, clone: EPR6855, abcam, 1/250 dilution, AR9 buffer, Opal 690) and IL17 (BS-2140R, ThermoFisher,1/400 dilution, AR6 buffer, Opal 520). DAPI was used as a nuclear counter stain. The stained TMA was scanned by fluorescence scanner (PANNORAMIC 250, 3DHISTECH). The cell annotation was performed by using QuPath.

#### LEGENDplex™ Cytokine assay

For protein analysis, the skin tissues from HC and DD patients were snap frozen at -70 °C until use. Skin tissues were lysed in PBS containing Protease Inhibitor Cocktail (1:100) (Sigma-Aldrich; P8340) with the final concentration of 16-64 mg skin/ml using innuSPEED Lysis Tubes J (Analytik Jena, Cat.: 845-CS-1120250) and SpeedMill PLUS (Analytik Jena). Lysate was filtered through 0.22 μm SpinX columns (Sigma; CLS8161) and analysed for cytokines using LEGENDplex™ HU Th (12-plex) assay (Biolegend, Cat.: 741028). Assay samples were acquired using Cytoflex LS (Beckman Coulter) flow cytometer. Data analysis was done using LEGENDplex™ Data Analysis Software Suite (QOGNIT).

#### Multi-color Flow Cytometry

For skin flow cytometry staining, biopsies were thawed, weighed and minced into smaller pieces using surgical scissors and were digested overnight with 1 ml skin digestion mix containing collagenase type 4 (0.8mg/ml; Worthington, LS004186) and DNAse (11.77 Units/ml; Sigma-Aldrich, D5319) in RPMI-complete (RPMIc) for about 0.1 g of skin at 37°C in 5% CO_2_ incubator [RPMIc: RPMI 1640 (Gibco, 31870074) with 5% human serum (One lambda, A25761), 1% penicillin/streptomycin (Sigma-Aldrich, P0781), 1% l-glutamine (Gibco, A2916801), 1% non-essential amino acid solution (NEAA) (Gibco, 11140035), 1% sodium pyruvate (Sigma-Aldrich, S8636), and 0.1% β-mercaptoethanol (Gibco, 31350-010)]. After overnight digestion, the cell suspension was filtered through 70 μm mesh, then washed and resuspended in RPMIc. For detection of intracellular cytokines, skin single-cell suspensions were stimulated with phorbol 12-myristate 13-acetate (PMA) (50 ng/ml; Sigma-Aldrich, P8139) and ionomycin (1 μg/ml; Sigma-Aldrich, I06434) with brefeldin A (10 μg/ml; Sigma-Aldrich, B6542) for 3 hours at 37 °C in 5% CO_2_ incubator. Cells were stained in PBS for surface markers (using antibodies: anti-CD3 bv605 (SK7), Biolegend, 344836; anti-CD4 PE-Cy5 (RPA-T4), Biolegend, 300510; anti-CD45 bv510 (HI30), Biolegend, 304036), and fixed and permeabilized using Foxp3 staining kit (Invitrogen; 00-5523-00) for staining the intracellular markers (using antibodies: anti-IL17A BV786 (N49-653), BD, 563745; anti-IL-22 PE (22URTI), eBioscience, 12-7229-42).

For PBMCs flow cytometry, cryopreserved PBMCs were thawed and rested overnight in RPMIc at 37°C in 5% CO_2_ incubator. Cells were then stimulated with PMA/ionocycin/brefeldin A as described above and stained in PBS for surface markers (using antibodies: anti-CD3 PE-Cy5 (UCHT1), Biolegend, 300410; anti-CD3 bv421 (UCHT1), Biolegend, 300434; anti-CD4 PE/Dazzle594 (RPA-T4), Biolegend, 300548; anti-CD4 PE-Cy5 (RPA-T4), Biolegend, 300510; anti-CD45RA AF700 (HI100), Biolegend, 304120; anti-CLA bv605 (HECA-452), BD, 563960; anti-CD103 APC (BerACT8), Biolegend, 350216). Cells were fixed and permeabilized using Cytofix/Cytoperm kit (Becton Dickinson, RUO 554714) for detection of intracellular IL-17A cytokine (using antibody: anti-IL-17A PerCPcy5.5 (BL168), Biolegend, 512314).

Data was acquired on Cytoflex LS (Beckman Coulter) flow cytometer and analyzed using FlowJo software (FlowJo LLC).

## Data Availability

All data produced in the present study are available upon request to the authors.
NanoString and scRNAseq data are available online at GSE222043, GSE235255.

## Data availability

The source data of this manuscript are provided as a Source Data file and are deposited in open public repositories (<u>GSE222043</u>, GSE235255). The scRNA-seq of healthy and psoriasis data used is from publicly available dataset (GSE162183).

## Code availability

The code used for scRNA-seq analysis is accessible in an online repository (https://github.com/GuenovaLab/Darrier).

## Acknowledgements

We would like to thank the patients who supported this research project. We would also like to thank all the physicians and nurses who cared for the patients. We thank Alessandra Darbellay and Ionoss Tabib for excellent technical support. We thank Dr. Antonia Currie for proofreading the manuscript. This work was supported by grants of the Medical Faculty of the Johannes Kepler University (Linz, Austria), the Swiss National Science Foundation (IZLIZ3_200253/1), the University of Lausanne (SKINTEGRITY.CH collaborative research program), and the Forschungskredit of the University of Zurich (FK-15-040 to WH).

## Author contributions

ME, SK and WH planned the study, revised the data and wrote the manuscript. TB processed samples from the clinics, performed qPCR, NanoString and Immunofluorecence experiments, YTC and YCT performed the IL-17 staining and the scRNA-seq experiment, AS performed FACS and bead-based cytokine detection experiments, PP was responsible for the scRNA-seq analysis, SRV contributed to performance and analysis of FACS and bead-based cytokine detection experiments, LS established and performed bead-based cytokine detection experiments, CI performed TMA assembly and analysis, ME, IP, JT and GW provided samples of patients and controls and clinical data, AL processed clinical samples and performed qPCR. SK analyzed the data and assembled the figures. ME, IG, SA, EG and WH revised the data and the manuscript.

## References

1. Sakuntabhai, A. et al. Mutations in ATP2A2, encoding a Ca2+ pump, cause Darier disease. Nat Genet 21, 271–277 (1999).

2. Cederlöf, M. et al. Darier disease is associated with type 1 diabetes: Findings from a population-based cohort study. J. Am. Acad. Dermatol. 81, 1425–1426 (2019).

3. Bachar-Wikström, E. & Wikström, J. D. Darier disease – a multi-organ condition? Acta Derm. Venereol. 101, (2021).

4. Dodiuk-Gad, R. P. et al. Darier disease in Israel: Combined evaluation of genetic and neuropsychiatric aspects. Br. J. Dermatol. 174, 562–568 (2016).

5. Bachar-Wikstrom, E. et al. Darier disease is associated with heart failure: a cross-sectional case-control and population based study. Sci. Rep. 10, 1–8 (2020).

6. Dodiuk-Gad, R. et al. Bacteriological aspects of Darier’s disease. J. Eur. Acad. Dermatology Venereol. 27, 1405–1409 (2013).

7. Curman, P. et al. Patients with darier disease exhibit cognitive impairment while patients with hailey-hailey disease do not: An experimental, matched case-control study. Acta Derm. Venereol. 101, 1–6 (2021).

8. Cederlöf, M. et al. Intellectual disability and cognitive ability in Darier disease: Swedish nation-wide study. Br. J. Dermatol. 173, 155–158 (2015).

9. Rogner, D. F., Lammer, J., Zink, A. & Hamm, H. Darier and Hailey-Hailey disease: update 2021. JDDG-J. Ger. Soc. Dermatology 19, 1478–1501 (2021).

10. Hanna, N., Lam, M., Fleming, P. & Lynde, C. W. Therapeutic Options for the Treatment of Darier’s Disease: A Comprehensive Review of the Literature. J. Cutan. Med. Surg. 26, 280–290 (2022).

11. Dicken, C. H. et al. Isotretinoin treatment of Darier’s disease. J. Am. Acad. Dermatol. 6, 721–726 (1982).

12. Sugiura, K. Unfolded protein response in keratinocytes: Impact on normal and abnormal keratinization. J. Dermatol. Sci. 69, 181–186 (2013).

13. Park, K., Lee, S. E., Shin, K. O. & Uchida, Y. Insights into the role of endoplasmic reticulum stress in skin function and associated diseases. FEBS J. 286, 413–425 (2019).

14. Li, N., Park, M., Xiao, S., Liu, Z. & Diaz, L. A. ER-to-Golgi blockade of nascent desmosomal cadherins in SERCA2-inhibited keratinocytes: Implications for Darier’s disease. Traffic 18, 232–241 (2017).

15. Chu, E. Y. et al. Diverse cutaneous side effects associated with BRAF inhibitor therapy: a clinicopathologic study. J Am Acad Dermatol 67, 1265–1272 (2012).

16. Savignac, M., Edir, A., Simon, M. & Hovnanian, A. Darier disease: A disease model of impaired calcium homeostasis in the skin. Biochim. Biophys. Acta - Mol. Cell Res. 1813, 1111–1117 (2011).

17. Gao, Y. et al. Single cell transcriptional zonation of human psoriasis skin identifies an alternative immunoregulatory axis conducted by skin resident cells. Cell Death Dis. 12, (2021).

18. Lowes, M. A. et al. Psoriasis Vulgaris Lesions Contain Discrete Populations of Th1 and Th17 T Cells. J. Invest. Dermatol. 128, 1207–1211 (2008).

19. Schnell, A., Littman, D. R. & Kuchroo, V. K. TH17 cell heterogeneity and its role in tissue inflammation. Nat. Immunol. 24, 19–29 (2023).

20. Werfel, T. et al. Cellular and molecular immunologic mechanisms in patients with atopic dermatitis. J. Allergy Clin. Immunol. 138, 336–349 (2016).

21. Klicznik, M. M. et al. Human CD4 + CD103 + cutaneous resident memory T cells are found in the circulation of healthy individuals. Sci. Immunol. 4, eaav8995 (2019).

22. Morren, M. A. et al. Challenges in Treating Genodermatoses: New Therapies at the Horizon. Front. Pharmacol. 12, 1–25 (2022).

23. Chen, C. et al. Sarco/endoplasmic reticulum Ca2+-ATPase (SERCA) activity is required for V(D)J recombination. 218, (2021).

24. Saluzzo, S. et al. Processing human skin samples for single-cell assays. STAR Protoc. 3, 101470 (2022).

25. Subramanian, A. et al. Gene set enrichment analysis: A knowledge-based approach for interpreting genome-wide expression profiles. Proc. Natl. Acad. Sci. U. S. A. 102, 15545–15550 (2005).

26. R_Core_Team. R: A language and environment for statistical computing. (2022).

27. Barter, R. L., Yu, B., Barter, R. L. & Yu, B. Superheat : An R Package for Creating Beautiful and Extendable Heatmaps for Visualizing Complex Data. 8600, (2018).

28. Hao, Y. et al. Integrated analysis of multimodal single-cell data. Cell 184, 3573-3587.e29 (2021).

29. Korsunsky, I. et al. Fast, sensitive and accurate integration of single-cell data with Harmony. Nat. Methods 16, 1289–1296 (2019).

