## Supplementary material for "Targeting of Th17-related cytokines in patients with Darier Disease": Suppl.

### Supplementary Information

#### Supplementary Tables

***Supplementary Table 1: Enriched pathways in keratinocytes of DD patients vs. Healthy Controls as determined by Gene Set Enrichment Analysis of scRNA-seq data.***

| Pathway name | Overlap | P value | Adjusted p value | database |
|---|---|---|---|---|
| mTORC1 Signaling | 9/200 | 0.00024 | 0.00876 | MSigDB_Hallmark_2020 |
| p53 Pathway | 8/200 | 0.00113 | 0.0209 | MSigDB_Hallmark_2020 |
| Toll-like receptor binding (GO:0035325) | 3/11 | 0.00017 | 0.04531 | GO_Molecular_Function_2021 |
| calcium ion binding (GO:0005509) | 11/348 | 0.00101 | 0.06961 | GO_Molecular_Function_2021 |
| RNA binding (GO:0003723) | 27/1406 | 0.0013 | 0.06961 | GO_Molecular_Function_2021 |
| icosatetraenoic acid binding (GO:0050543) | 2/6 | 0.00154 | 0.06961 | GO_Molecular_Function_2021 |
| arachidonic acid binding (GO:0050544) | 2/6 | 0.00154 | 0.06961 | GO_Molecular_Function_2021 |
| gap junction channel activity involved in cell communication by electrical coupling (GO:1903763) | 2/6 | 0.00154 | 0.06961 | GO_Molecular_Function_2021 |
| icosanoid binding (GO:0050542) | 2/7 | 0.00214 | 0.07259 | GO_Molecular_Function_2021 |
| insulin-like growth factor II binding (GO:0031995) | 2/7 | 0.00214 | 0.07259 | GO_Molecular_Function_2021 |
| metal ion binding (GO:0046872) | 13/517 | 0.00282 | 0.08485 | GO_Molecular_Function_2021 |
| snoRNA binding (GO:0030515) | 3/30 | 0.00356 | 0.08928 | GO_Molecular_Function_2021 |
| RAGE receptor binding (GO:0050786) | 2/9 | 0.00362 | 0.08928 | GO_Molecular_Function_2021 |
| RB DN.V1 DN | 9/126 | 6.4E-06 | 0.00099 | MSigDB_Oncogenic_Signatures |

***Supplementary Table 2: Enriched Biocarta pathway gene sets in Darier Disease patients vs. Healthy Controls as determined by Gene Set Enrichment Analysis of NanoString data.***

IL-17 signaling pathway is highlighted in bold. FDR, false discovery rate

| Biocarta pathway name | No of genes in Nanostring panel | normalized enrichment score | FDR q-value |
|---|---|---|---|
| Cytokines and inflammatory response | 21 | 2.1 | 0.05 |
| Local acute inflammatory response | 15 | 2.05 | 0.07 |
| Adhesion and diapedesis of granulocytes | 13 | 2.03 | 0.074 |
| **IL-17 signaling** | 12 | 1.98 | 0.124 |
| Cytokine network | 17 | 1.86 | 0.193 |
| Cytotoxic T lymphocytes | 11 | 1.84 | 0.196 |
| Regulation of hematopoiesis by cytokines | 12 | 1.81 | 0.18 |
| NFkB activation by nontypeable Hemophilus influenzae | 14 | 1.75 | 0.196 |
| T helper cells | 10 | 1.71 | 0.225 |
| Adhesion and diapedesis of lymphocytes | 12 | 1.71 | 0.207 |
| T cytotoxic cells | 10 | 1.68 | 0.219 |
| Free radical induced apoptosis | 4 | 1.66 | 0.216 |
| Adhesion molecules on lymphocytes | 9 | 1.61 | 0.217 |
| HIV induced T cell apoptosis | 7 | 1.62 | 0.221 |
| Co-stimulatory signal during T-cell activation | 13 | 1.6 | 0.212 |
| Monocytes and its surface molecules | 10 | 1.56 | 0.226 |

***Supplementary Table 3: Enriched Biocarta pathway gene sets in Psoriasis patients vs. Healthy Controls as determined by Gene Set Enrichment Analysis of NanoString data.***

Pathways that are also enriched in Darier Disease patients are highlighted in bold. FDR, false discovery rate

| Biocarta pathway name | No of genes in Nanostring panel | normalized enrichment score | FDR q-value |
|---|---|---|---|
| **IL-17 signaling** | 12 | 2.32 | 0.174 |
| **Cytokine network** | 17 | 2.27 | 0.174 |
| **Regulation of hematopoiesis by cytokines** | 12 | 2.11 | 0.174 |
| **NFkB activation by nontypeable Hemophilus influenzae** | 14 | 2.06 | 0.174 |
| **Cytokines and inflammatory response** | 21 | 1.82 | 0.174 |
| **Free radical Induced apoptosis** | 4 | 1.82 | 0.174 |
| IL22 Soluble receptor signaling | 9 | 1.76 | 0.174 |
| **Adhesion and diapedesis of granulocytes** | 13 | 1.74 | 0.174 |
| Mechanism of gene regulation by peroxisome proliferators via PPARa | 8 | 1.72 | 0.174 |
| **Local acute inflammatory response** | 15 | 1.6 | 0.224 |
| **Adhesion and diapedesis of lymphocytes** | 12 | 1.59 | 0.22 |
| Antigen processing and presentation | 11 | 1.52 | 0.231 |

#### Supplementary Figures

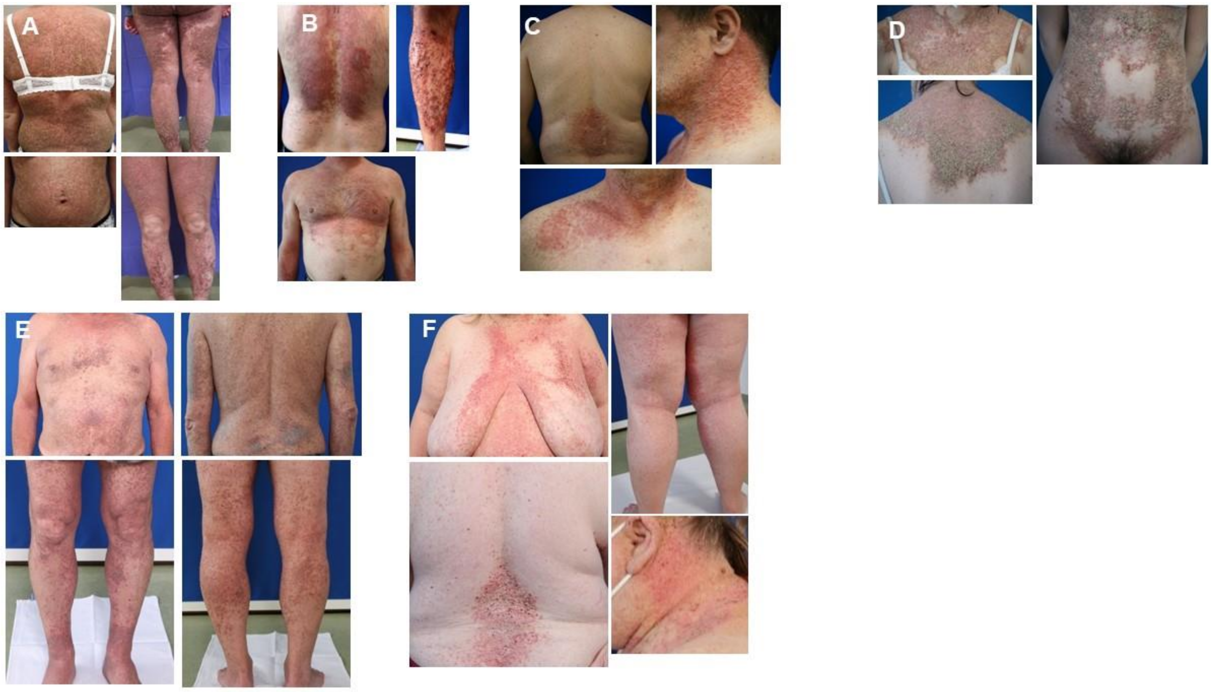

***Supplementary Fig 1: Disease phenotype/severity***

**A** PAT1: hyperkeratotic erythematous and brownish papules and plaques on the entire integument, on legs additionally erosions and crusts (severe, >35% BSA affected). **B** PAT2: hyperkeratotic erythematous and brownish papules and plaques on the entire integument, on the legs additionally erosions and crusts (severe, >35% BSA affected). **C** PAT4: hyperkeratotic erythematous and brownish papules and plaques on the upper thorax and neck (moderate, 20% BSA affected). **D** PAT5: hyperkeratotic erythematous and brownish papules and plaques on the entire integument (severe, >35% BSA affected). **E** PAT7 hyperkeratotic erythematous and brownish papules and plaques on the entire integument (severe, >35% BSA affected). **F** PAT9: hyperkeratotic erythematous and brownish papules and plaques on the entire integument (severe, >35% BSA affected). BSA, body surface area

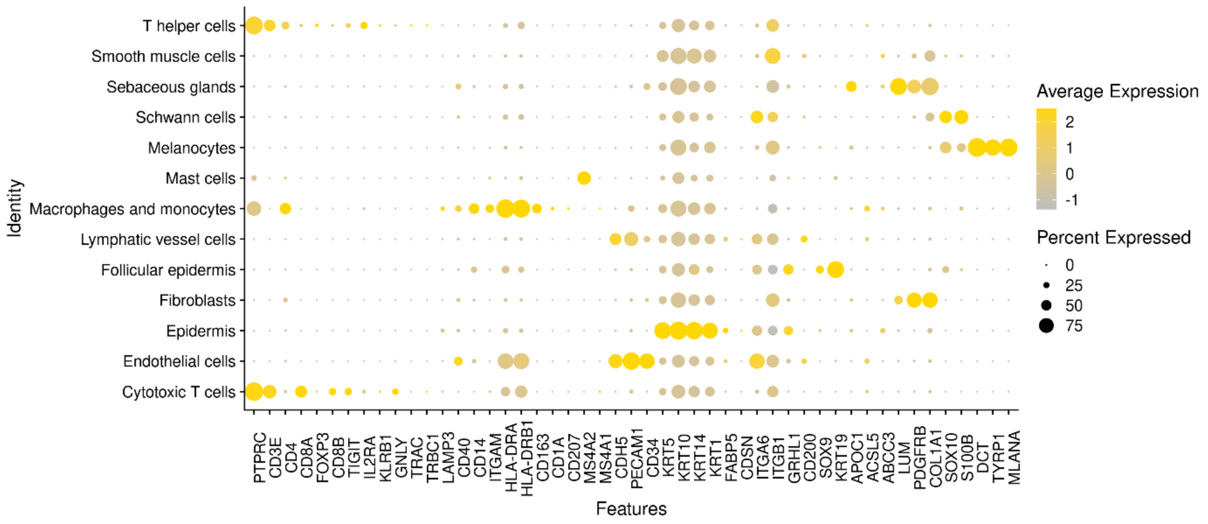

***Supplementary Fig 2: scRNA-seq characterization of cell types***

Dot plot demonstrating mean log normalized expression of cell type-specific marker genes. Color scale indicates mean expression, and circle size indicates fraction of cells in a group.

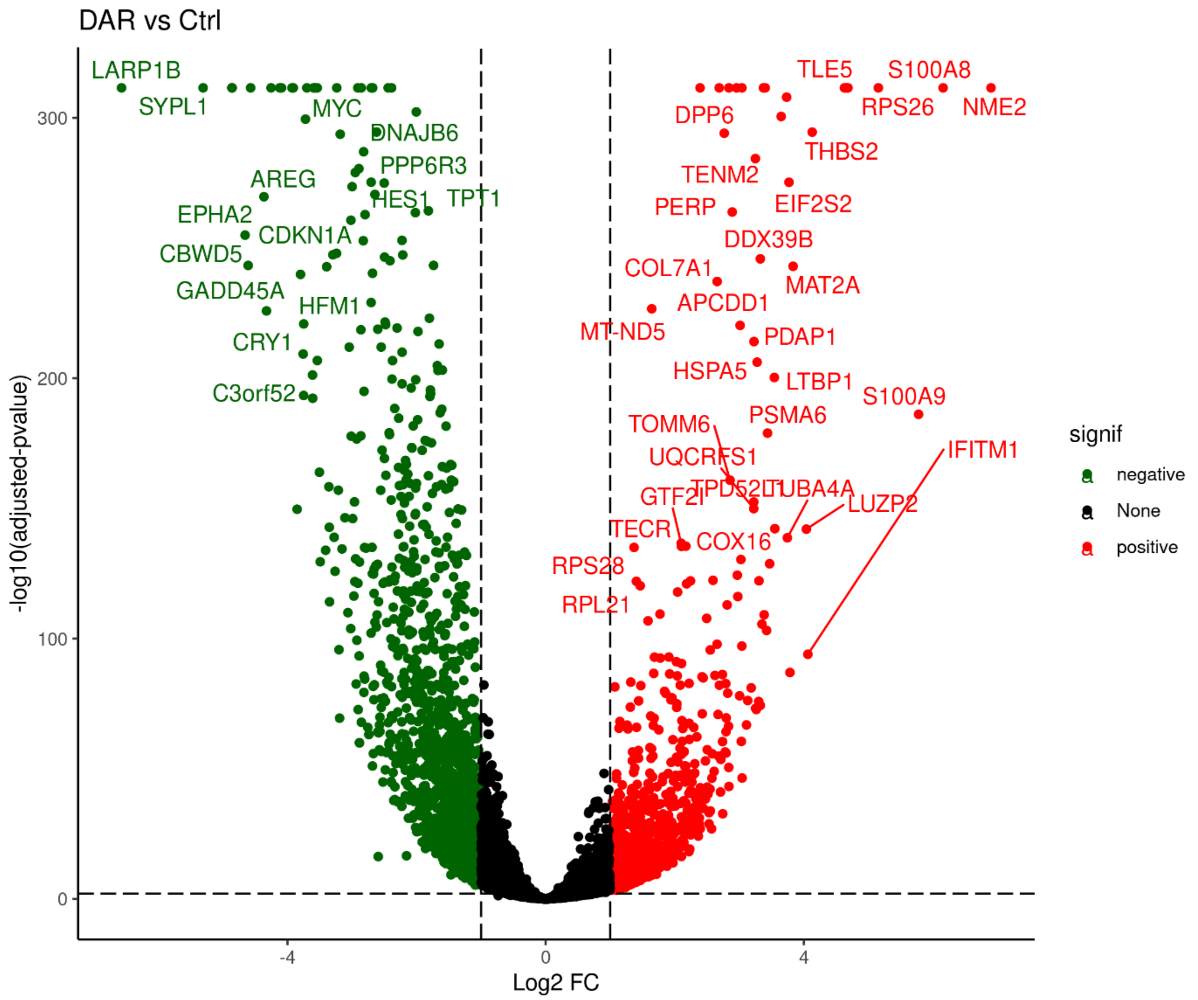

***Supplementary Fig 3: Volcano plot of differentially expressed genes in keratinocytes of DD patients vs HC***

scRNA-seq revealed a plethora of DE genes in keratinocytes of DD patients as compared to healthy control skin.

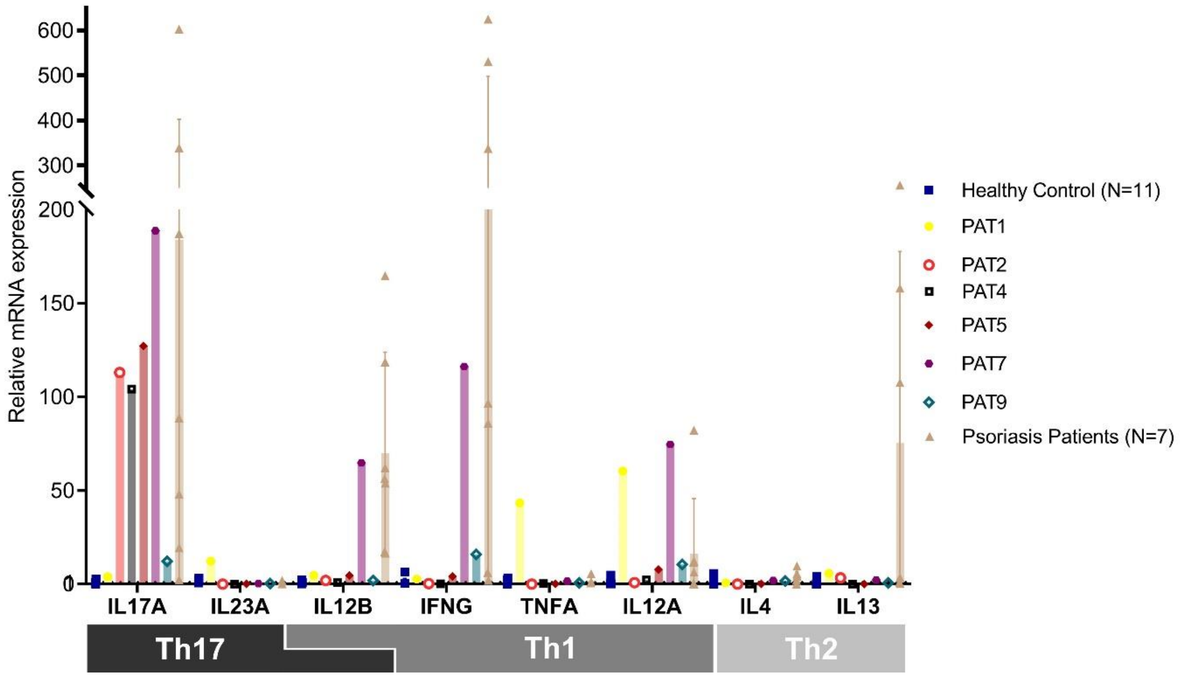

***Supplementary Fig 4: Cytokine mRNA expression of each single patient***

Cytokine mRNA expression of each single patient was determined by qPCR using probes against the indicated cytokines. Data were normalized to housekeeping gene ACTB and healthy controls. Data points represent means of individuals. Bars represent means, error bars represent standard deviations.

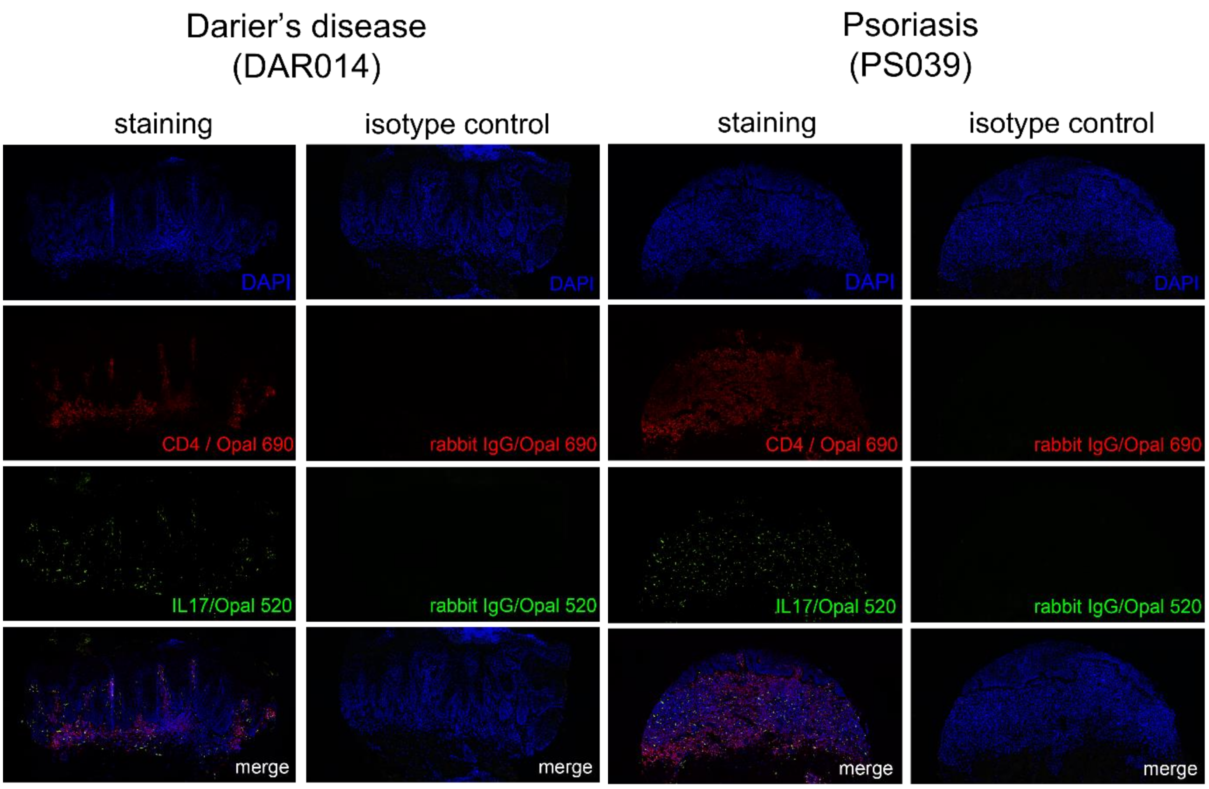

***Supplementary Fig 5: Isotype controls of multi-color IF images***

Left panel is one representative DD patient, right panel is one representative PSO patient.

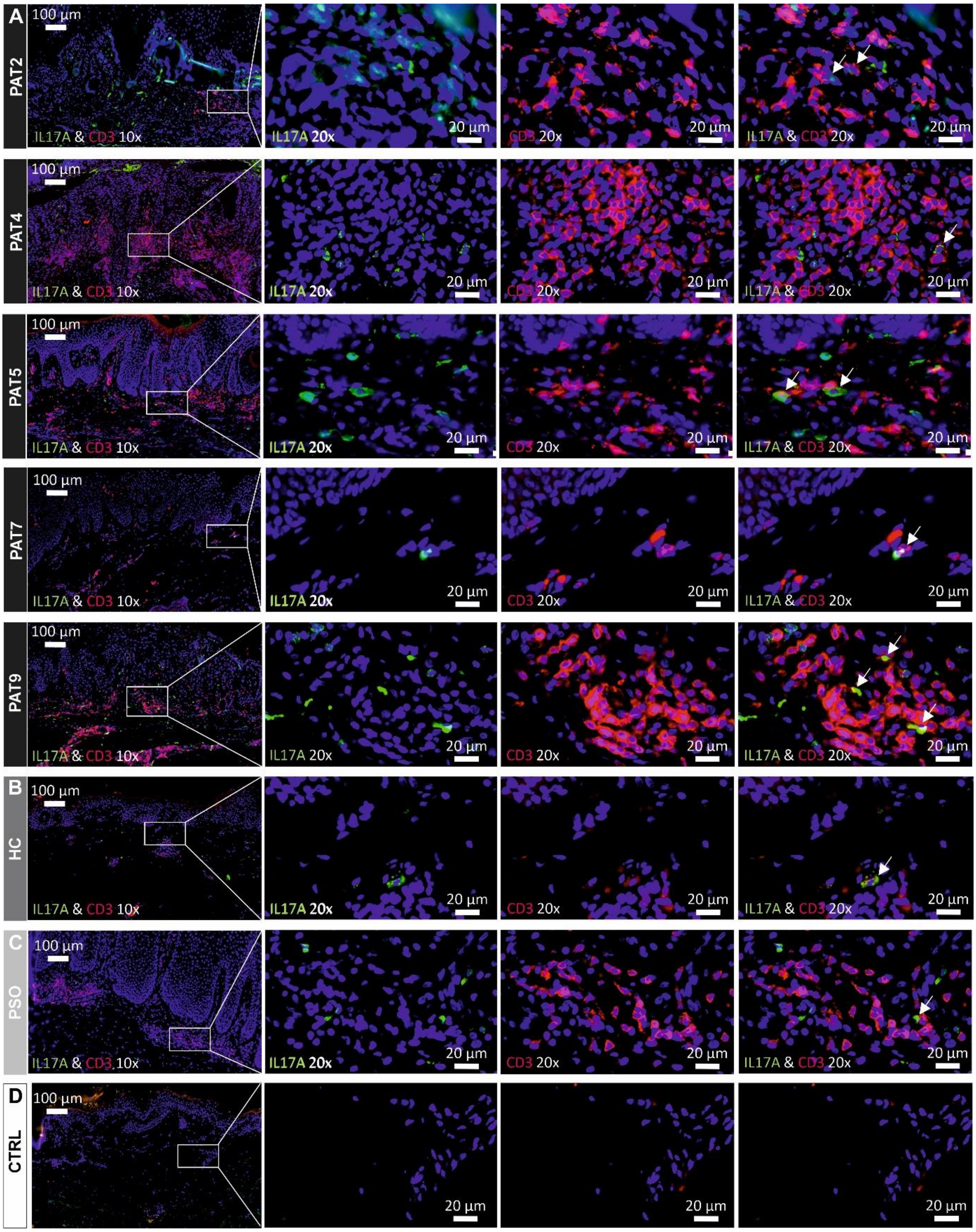

***Supplementary Fig 6: Immunofluorescence stainings of IL17A and CD3***

**A** Skin sections of DD patients (PAT2-9) were stained with antibodies against IL17A and CD3. **B** Immunofluorescence images of IL17A$^{+}$CD3$^{+}$ from healthy control (HC). **C** Immunofluorescence images IL17A$^{+}$CD3$^{+}$ of a psoriasis patient (PSO). CD3 positive T cells in red, IL17A positive cells in green, DAPI-stained nuclei in blue. White arrows indicate IL17A/CD3 double-positive cells (supposedly Th17/Tc17 cells) in 20x magnified images. **D** Secondary AB control

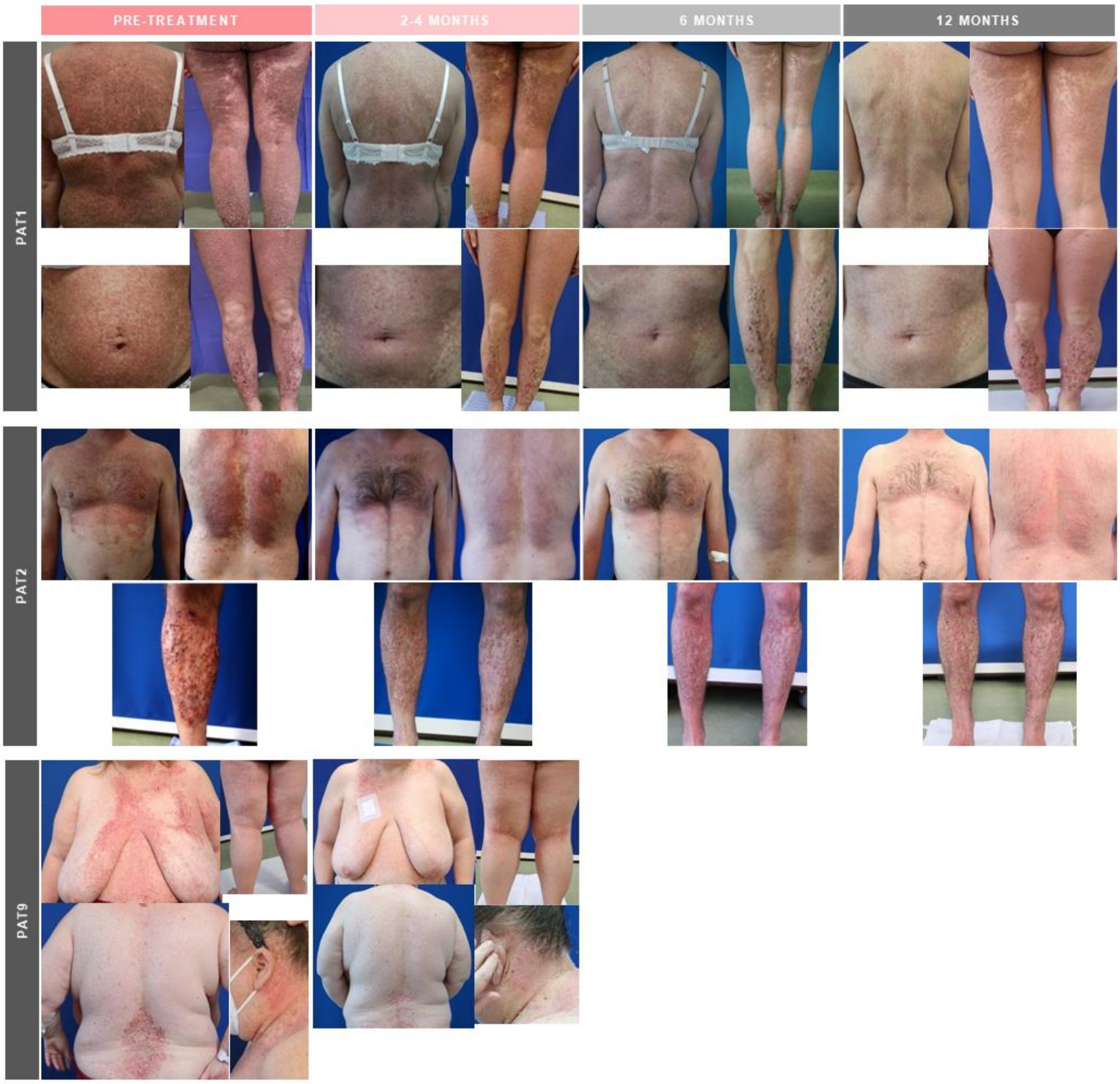

***Supplementary Fig 7: Clinical pictures during treatment of DD PAT1, PAT2 and PAT9***

**Patient 1:** Pre-treatment (prior to systemic therapy with guselkumab): erythematous and brownish hyperkeratotic papules and plaques on the entire integument, on the legs additionally crusts and erosions. 2 months post therapy: flattening of papules and plaques on the thorax, decrease of erythema. 6 months post therapy: further flattening of papules and plaques on the thorax. 12 months post treatment: flattened and reduced papules on the thorax, on the legs hyperkeratotic plaques and erythema, crusts and erosions.

**Patient 2**: Pre-treatment (before systemic therapy with secukinumab): erythematous and brownish hyperkeratotic papules and plaques on thorax and legs, on legs additionally crusts and erosions. 3 months post therapy: reduced erythema on the thorax, flattened papules on the thorax and plaques on the legs. 6 months post therapy: flattened papules on the thorax. 12 months post therapy: flattened and reduced papules on the thorax, hyperkeratotic plaques on the legs and lightened erythema.

**Patient 9:** Pre-treatment (prior to systemic therapy with secukinumab): erythematous and brownish hyperkeratotic papules and plaques on thorax, legs and face. 3.5 months post therapy: flattened plaques on thorax and legs.

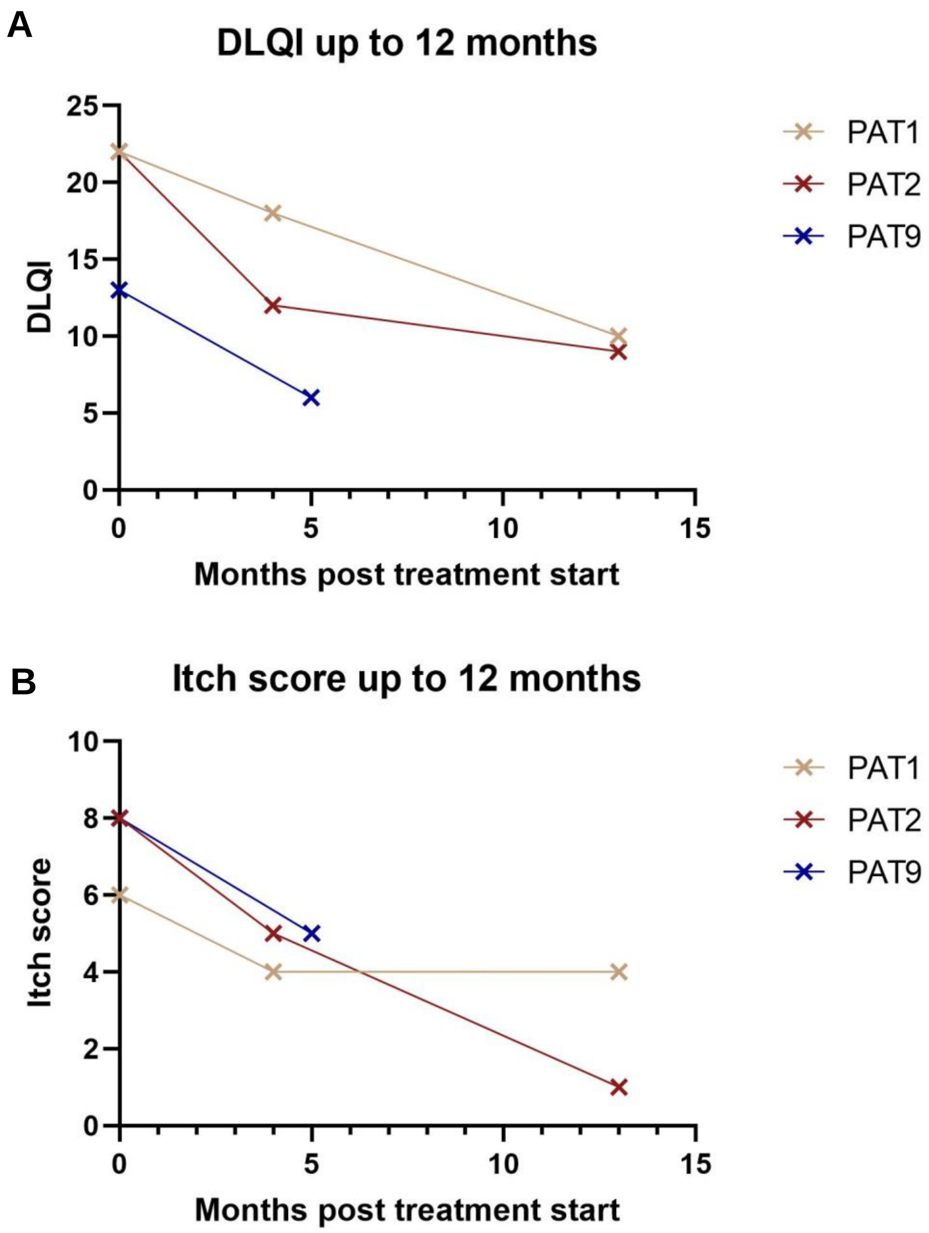

***Supplementary Fig 8: Improved itch and quality of life of DD patients treated with biologics.***

**A** Dermatology Life Quality Index questionnaire (DLQI) score of PAT1, PAT2 and PAT9. **B** Itch score (visual analogue scale 0-10) of PAT1, PAT2 and PAT9. Both scores were retrospectively collected from PAT1 and PAT2, while they were prospectively collected from PAT9.

#### Supplementary Methods

##### *Immunofluorescence.*

Immunofluorescence was performed on formalin-fixed, paraffin-embedded lesional patient skin and normal control skin. After deparaffinization of the slides, Antigen Retrieval was performed in boiling hot citrate buffer (pH 6.0) for ten minutes. Slides were blocked, followed by overnight incubation of the 1:500 diluted IL-23 p19 Rabbit anti-Human Antibody (PA520239, Invitrogen), 1:200 diluted IL-17 Polyclonal Antibody (bs-2140R, Bioss) and 1:50 diluted Anti-Human CD3 Monoclonal Antibody (MA5-12577, Invitrogen). Antibodies Goat anti-Rabbit IgG (H+L) Cross-Adsorbed Secondary Antibody, AF 488 (A-11070, Thermo Fisher Scientific) and Donkey anti-Mouse IgG (H+L) Highly Cross-Adsorbed Secondary Antibody, AF 647 (A-31571, Invitrogen) were utilized as secondary antibodies. Slides were covered using Antifade/DAPI covering media and scanned with Histo Scanner VS200 v4 (Olympus).
